# Platinum vs non-platinum chemotherapy for platinum-resistant ovarian cancer: a systematic review and meta-analysis

**DOI:** 10.1101/2022.07.12.22277568

**Authors:** Alexey Rumyantsev, Alexandra Tyulyandina, Mikhail Fedyanin, Ilya Pokataev, Elena Glazkova, Sergey Tjulandin

## Abstract

**Background:** recurrent ovarian cancer (OC) patients with platinum-free interval (PFI) <6 mo. are usually considered platinum-resistant and treated with non-platinum based chemotherapy. However, this was never confirmed in proper-conducted randomized trials.

**Methods:** we queried the PubMed database for all full-text articles and abstracts on the treatment of patients with platinum-resistant ovarian cancer (PROC) in 01/01/2000-01/06/2019 timeframe. The PRISMA tool was used to ensure transparent reporting of the results. Inclusion criteria were: 1) morphologically confirmed epithelial ovarian cancer; 2) recurrent disease within 6 months after completion of platinum-based chemotherapy; 3) treatment with platinum- or non-platinum chemotherapy with agents that are routinely used for OC; 4) no concomitant therapy with targeted or investigational agents or non-platinum doublets; 5) defined response rate (RR) and assessment criteria. Proportion meta-analysis (random-effect model) and beta-regression were conducted to assess the impact of platinum agents on response rate as well as significance of other variables. In the beta-regression model response rate was a dependent variable, while platinum agents (yes or no), used non-platinum drugs and method of response assessment were independent variables. Statistical analysis was dose with meta, metafor and betareg packages of R software.

**Results:** we identified 7156 articles and screened them for title and abstract, 157 studies for further analysis. Overall, 6327 patients were included in the analysis, efficacy of non-platinum- and platinum-based therapy was assessed in 113 (n = 5272) and 44 (n = 1055) trials respectively. In meta-proportion random-effect model RR among patients treated with platinum-based and non-platinum chemotherapy RR was 36% (95% CI 30-41; I2 = 62%) and 16% (95% CI 14-19; I2=70%) respectively. For sensitivity analysis various regression models were made with different subsets of the trials and additional variables (including year of the trial, percentage of serous subtype of OC and median of prior therapy lines). Platinum was the strongest predictor of response in every developed model.

**Conclusion:** this meta-analysis shows that patients with ‘platinum-resistant’ ovarian carcinoma may derive significant benefit from reintroduction of platinum agents. These results support recent ESMO-ESGO consensus on treatment of recurrent ovarian cancer.

## Introduction

Recurrent ovarian cancer (ROC) is an incurable disease and treatment goals for these patients are disease control and maintaining quality of life. Durable remissions are linked to better survival and outcomes. Platinum agents are therapeutic backbone for advanced ovarian cancer and valuable drugs in ROC setting. The management of the disease is based on platinum-free interval (PFI), defined as time between platinum-based treatment competition and relapse. PFI guides treatment decisions as it tightly correlates with survival and response to subsequent anticancer treatment. Conventionally recurrent ovarian cancer is divided into two main categories^1^:

- Platinum-sensitive disease with PFI of six months or longer, these patients are deemed to have chemotherapy-sensitive disease and surely benefit from further platinum agents;
- Platinum-resistant disease with PFI of less than 6 months, these patients are considered to have disease that will not respond to subsequent platinum-based chemotherapy. Those patients whose disease progressed during platinum-based chemotherapy or within 4 weeks of last dose of platinum are called platinum-refractory^1^. The latter category has dismal prognosis.

On the other hand, many authors reported response rate (RR) in 30-60% of patients with platinum-resistant ovarian cancer (PROC) after repeated platinum rechallenge (platinum administration in patients already treated with platinum salts). Median PFS and OS were 5-8 months and 10-18 months respectively^2–14^. We conducted a systematic review and meta-analysis of published trials to compare efficacy of platinum rechallenge versus non-platinum chemotherapy in patients with PROC.

## Materials and methods

### Search strategy

PubMed database queries have been performed with predefined criteria to include all prospective and retrospective full-text articles and abstracts for all years in 01/01/2000-01/07/2019 timeline in English. In the search queries, the criterion for the publication of clinical and observational human studies was used as an additional filter; surveys, guidelines and other publications that did not contain original data were excluded from the search. We used the following query algorithm in PubMed: (ovarian*[Title] OR OVARIAN NEOPLASMS [MESH]) AND (RESISTANT OR RECURRENT OR PLATINUM-RESISTANT OR REFRACTORY OR PLATINUM-REFRACTORY).

### Selection of the trials

All identified records were individually screened for eligibility on title and abstract by the two authors (RAA and TAS). Discrepancies were discussed and resolved by consensus, resulting in either inclusion or exclusion of the article. The Preferred Reporting Items for Systematic Reviews and Meta-Analyses (PRISMA) tool was used to ensure transparent reporting of the results. Study inclusion criteria were:

1. Morphologically confirmed epithelial ovarian cancer. We excluded publications that specifically addressed treatment of rare epithelial ovarian (ie, mucinous, clear cell or low-grade) cancers or non-epithelial (germ-cell and stromal) ovarian neoplasms. There were no limitations on inclusion of trials if patients with rare epithelial malignant ovarian tumors were treated as a subset of general population;
2. Recurrent ovarian cancer. For the purposes of this trial we adopted a standard definition of PROC as a disease that recurred within 6 months after completion of platinum-based chemotherapy. We excluded papers with any other cut-offs for PROC (ie, <12 months) or if no clear definition for PROC was provided by the authors;
3. Treatment with platinum-based or non-platinum chemotherapy with agents that are routinely used for ovarian cancer patients and listed in the NCCN guidelines as a possible options for patients with PROC, such as platinum compounds (cisplatin, carboplatin, oxaliplatin), taxanes (paclitaxel, docetaxel), anthracyclines (doxorubicin, epirubicin or pegylated lyposomal doxorubicin), antimetabolites (gemcitabine, capecitabine, 5-fluorouracil), topoisomerase inhibitors (etoposide, topotecan, irinotecan) etc^16^;
4. No concomitant therapy with targeted (including VEGF-targeting) or investigational agents, intraperitoneal or high-dose chemotherapy with stem cell support, non-platinum doublets;
5. Clearly defined response rate for patients with PROC and criteria used for response assessment (radiological, [i.e., RECIST or WHO/GOG criteria] vs CA-125-based response classification systems);
6. Availability of full text for the articles.

If the trial reported results for several cohorts separately (eg, randomized trials comparing two non-platinum agents or two dosing regimens) they were included as independent observations. In case of multiple publications based on the same study cohort we included the most relevant study (according to the above mentioned inclusion criteria). As the primary endpoint of the trial was response rate we excluded trials if no information about RR were available in the manuscript or supplementary files. We also excluded case reports, trials with single-agent platinum chemotherapy or with high-dose chemotherapy with stem-cell support, trials with investigational agents without standard-treatment control arms.

### Data analysis

This was a single-arm trials meta-analysis. We extracted the following information for every eligible trial from the included trials: year of publication, number of patients, regimen of chemotherapy, response rate (as raw numbers of responders and non-responders), response assessment criteria (CA-125 vs radiological response), median overall survival and median progression-free survival. Response rate was chosen as a primary endpoint for the analysis, hence meta-proportional meta-analysis was conducted with the mentioned outcome. The weights of individual studies were based on the inverse variance method, DerSimonian-Laird method was chosen as an estimator for tau^2^. Arcsine transformation method was selected for raw proportions. Meta-regression analysis was conducted to assess the role of various moderators on response rate.

Differences in trials characteristics between arms were assessed using general linear regression weighting trials according to number of enrolled patients. For the purposes of progression-free survival analysis we conducted a simple general linear regression analysis of log-transformed PFS treating each study included trial as an independent observation. The observations were weighted according to the number of enrolled patients. The association between the log-transformed median PFS time administration of platinum agents, various non-platinum agents, modality of response assessment and other factors was assessed. Multiple regression analysis was conducted to assess an impact of platinum-based chemotherapy of median PFS adjusting for other potential predictors.

All statistical analysis was done using R and RStudio statistical software.

## Results

In mentioned timeframe we identified 7156 articles and screened them for title and abstract. After the review process we selected 197 studies for further analysis. Of them, 44 (n = 1055) and 113 (n = 5272) trials assessed efficacy of platinum- and non-platinum based chemotherapy respectively. Only one randomized phase 2 trial^17^ was designed for direct comparison of platinum-based and non-platinum chemotherapy in PROC setting, the results of this trial were included into separate categories for the analysis. All other trials evaluating efficacy of platinum reintroduction in PROC patients were non-randomized phase 2 single-arm studies. The trials flowchart (PRISMA diagram) is provided in Figure 1. Overall 6327 patients were enrolled in the identified studies. Patients and trials characteristics are summarized in Table 1.

**Table 1.**
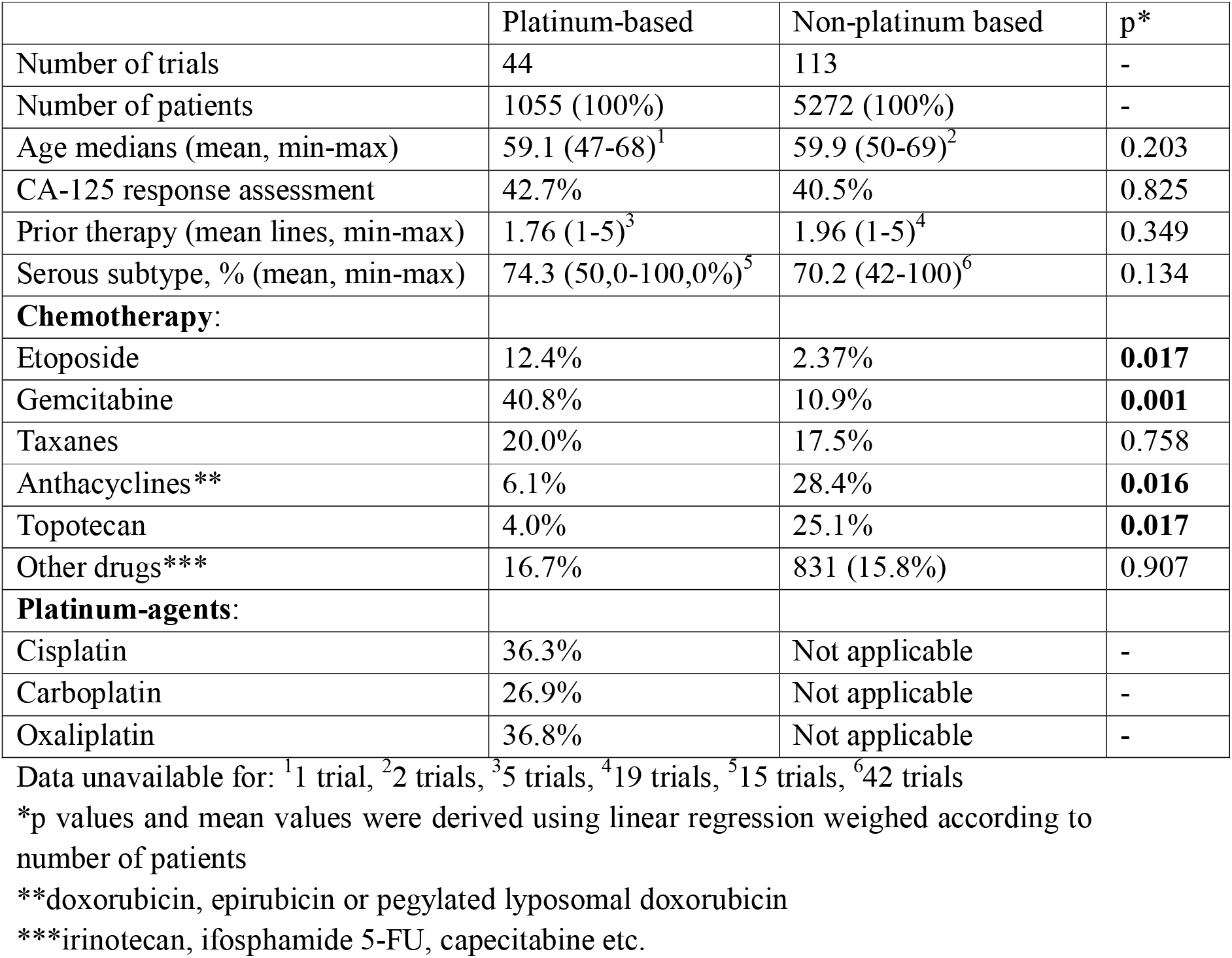
Patients & trials characteristics

**Figure 1.**
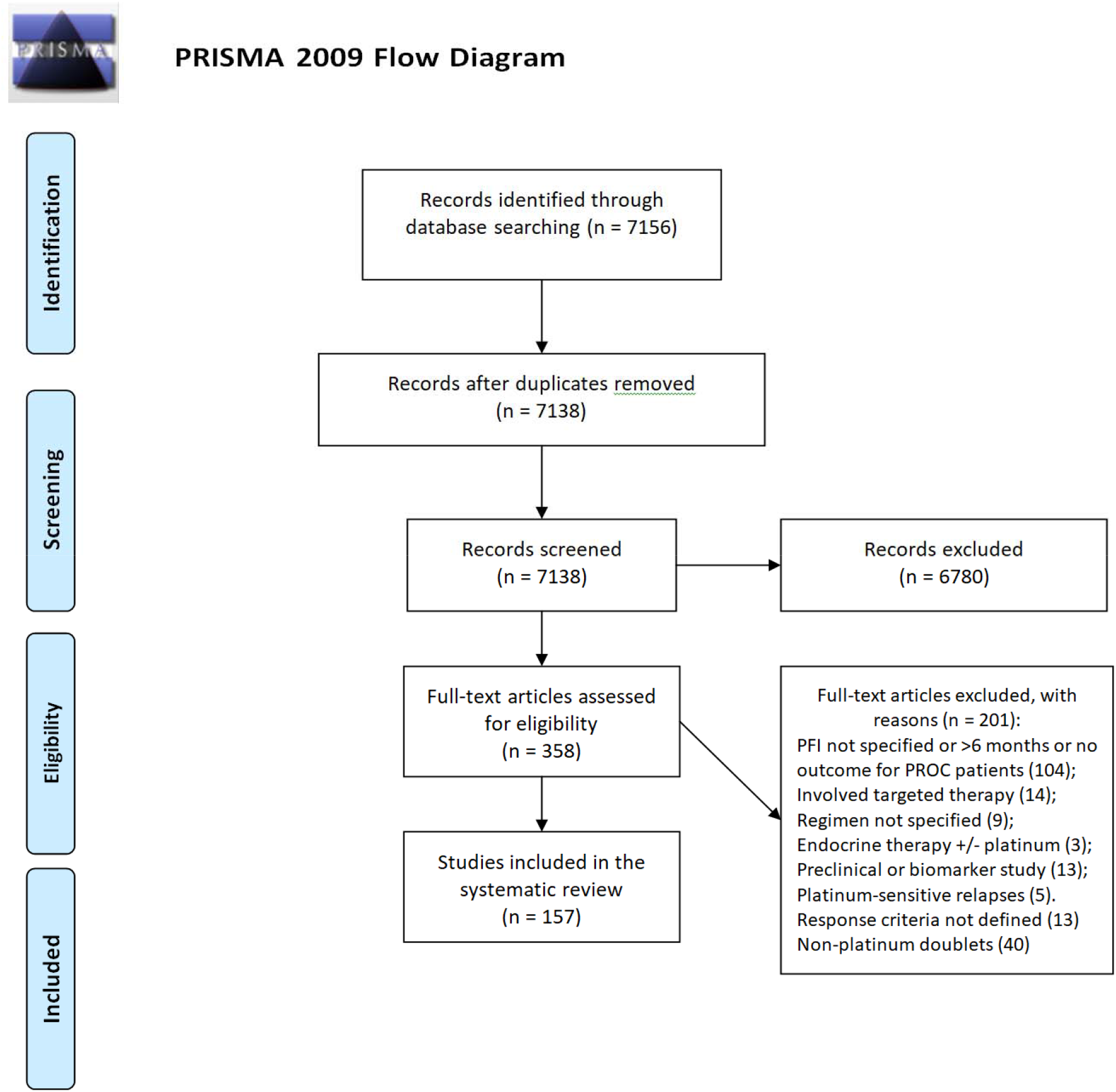
PRISM diagram for trial selection

### Platinum vs non-platinum based chemotherapy

6327 patients were included into this analysis, according to random-effects meta-analysis of proportions overall response rate in patients with platinum-resistant ovarian cancer was 21% (95% CI 0.19-0.24% [Tau^2^ = 0.03, Chi^2^ = 903.55, df = 156 (p <0.01), I^2^=83%]). Among patients treated with platinum-based and non-platinum chemotherapy objective response rate was 37.6% (95% CI 32.3-43.02 [I^2^=68%]) and 16.37% (95% CI 14.11-18.77 [I^2^=80%]) of patients respectively, test for subgroup differences was significant (Chi^2^=56.17, p < 0.01). Figure 2 depicts forest-plot for platinum-based chemotherapy trials (forest plot for non-platinum therapy available as a supplement to the article only due). The comprehensive list of trials that were included into analysis is available as supplement to this article.

**Figure 2.**
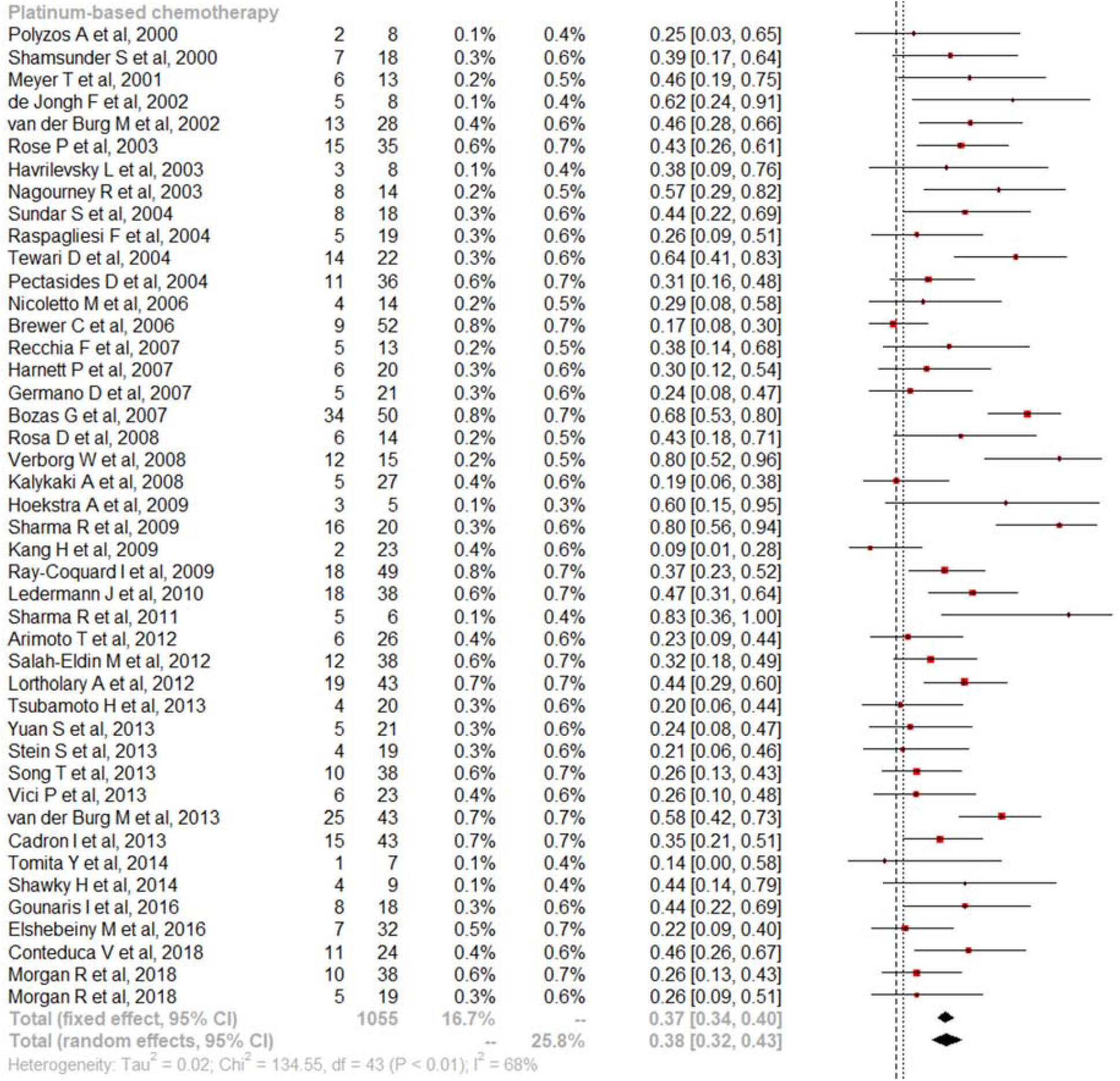
Forest plot for meta-proportion analysis of platinum-based therapy efficacy

To further evaluate the impact of platinum rechallenge on RR in PROC patients and address potential imbalances we performed meta-regression analysis. On the first step we conducted a univariable analysis to evaluate the probability of response under various factors (Table 2). We revealed that strongest predictor of response in PROC patients was administration of platinum agents with corresponding β_*1*_ value 0.243 (p <0.001). Biological models of response assessment, administration of taxanes and etoposide were other significant factors with positive impact on RR. Administration of topotecan inversely correlated with probability of response in unifactor analysis.

**Table 2.**
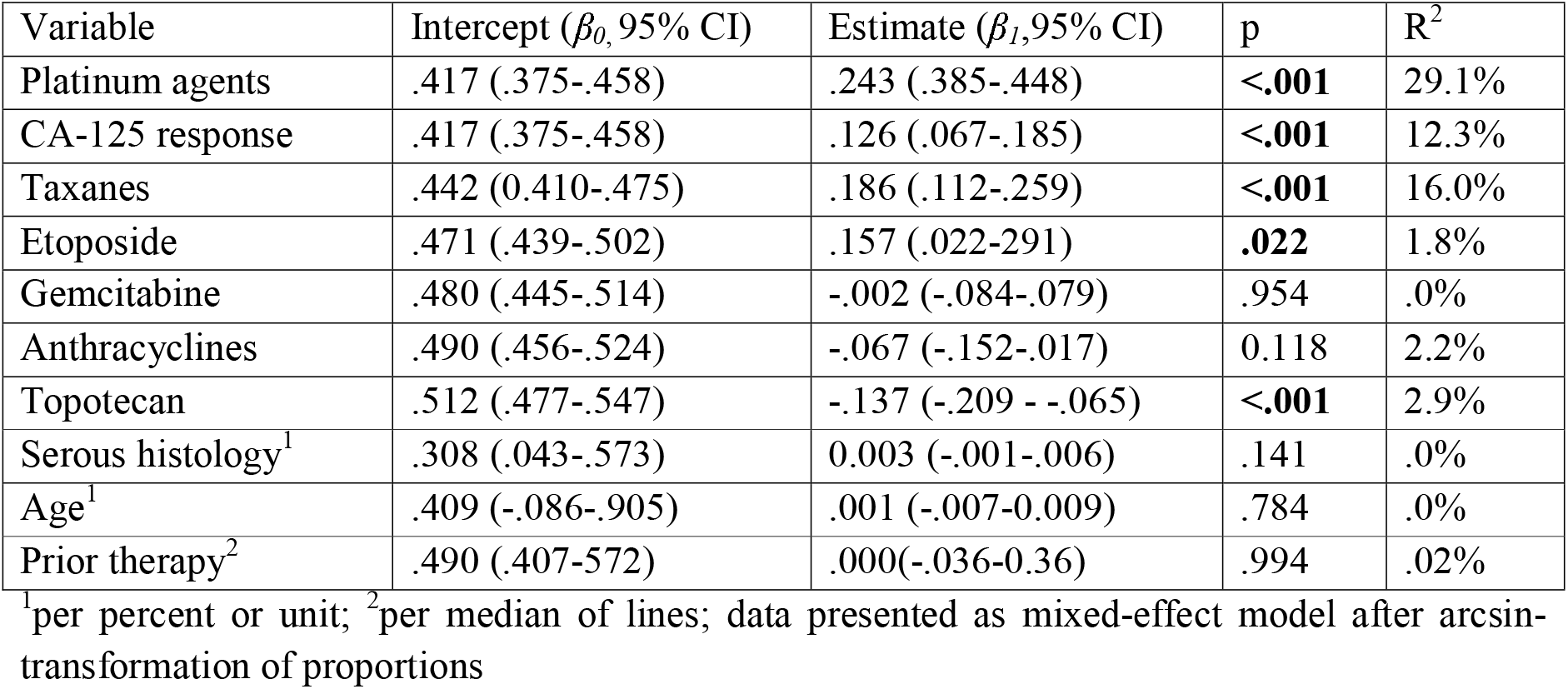
Meta-regression univariable analysis of variables impacting the probability of response

On the next step, we conducted a multifactor meta-regression analysis, including the factors significant in unifactor analysis. The results are summarized in Table 3. The model confirmed independent value of platinum chemotherapy as the strongest predictor of response even after adjustment for response assessment method and administered non-platinum agents. Among the latter, only administration of taxanes had an independent value and was associated with the best probability of response among other non-platinum agents, this was explored further. The mentioned values can be easily transformed to the probabilities, eg, under the model the predicted probability of response in PROC patient treated with platinum- and taxane-based chemotherapy is 44.37% compared to 9.83% for topotecan or other non-platinum monotherapy.

**Table 3.**
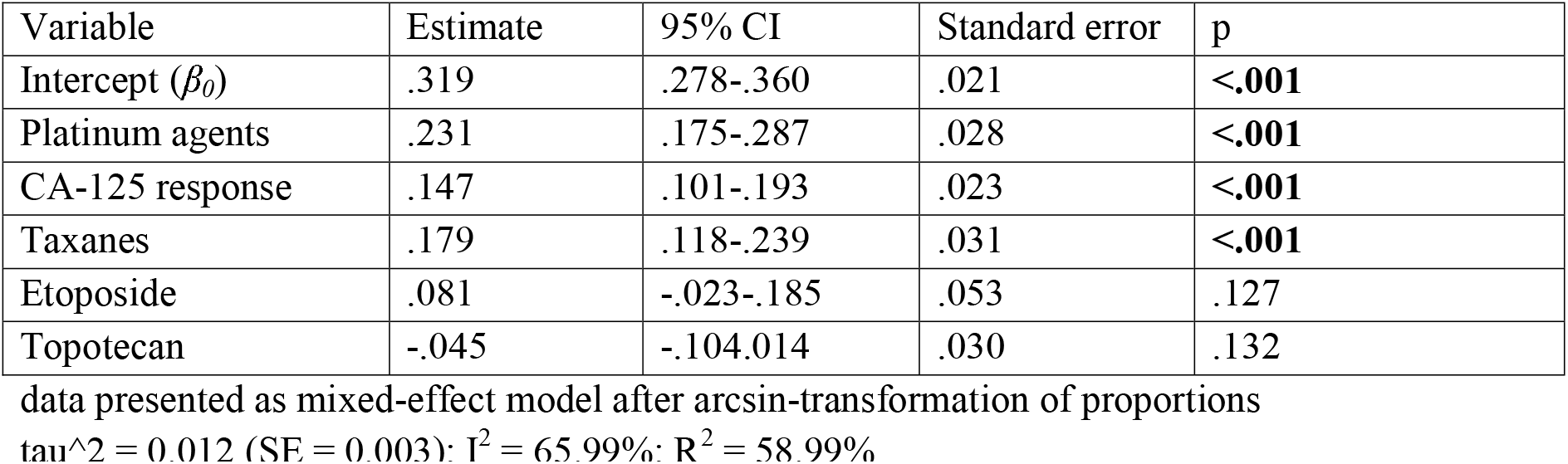
Multiple meta-regression model of variables impacting the probability of response

| Variable | Estimate | 95% CI | Standard error | p |
| --- | --- | --- | --- | --- |
| Intercept ($\beta_0$) | .319 | .278-.360 | .021 | **<.001** |
| Platinum agents | .231 | .175-.287 | .028 | **<.001** |
| CA-125 response | .147 | .101-.193 | .023 | **<.001** |
| Taxanes | .179 | .118-.239 | .031 | **<.001** |
| Etoposide | .081 | -.023-.185 | .053 | .127 |
| Topotecan | -.045 | -.104-.014 | .030 | .132 |

data presented as mixed-effect model after arcsin-transformation of proportions
$\tau^2 = 0.012$ (SE = 0.003); $I^2 = 65.99\%$ ; $R^2 = 58.99\%$

For sensitivity analysis purposes we explored several additional meta-analytical models, including the multivariate model with inclusion of all factors evaluated in unifactor analysis, mixed-effect multivariate meta-regression with random effects, generalized linear mixed-effect models (GLMM) beta-regression models, models with various raw proportions transformations (eg, logit, Freeman–Tukey double-arcsine transformation) and methods for estimation of the between-study variance. Every evaluated model returned essentially the same predictors and platinum agents were the most influential factor related response to chemotherapy.

We conducted a separate analysis to assess efficacy of various non-platinum agents in PROC setting (Table 4). For non-platinum drugs, taxanes were associated with the highest probability of response to anticancer treatment with estimated RR 22.5% (95% CI 15.2-30.9), while treatment with gemcitabine (6.8%; 95% CI 2.4-13.2) or topotecan (7.2%; 95% CI 2.8.5-13.3) had the lowest rates of objective response.

**Table 4.** Predicted efficacy of non-platinum agents as monotherapy in PROC

| Drug | Estimated RR | 95% CI | p* |
| --- | --- | --- | --- |
| Etoposide | 14.9% | 5.7-27.4% | .383 |
| Gemcitabine | 6.8% | 2.4-13.2% | .009 |
| Taxanes | 7.2% | 15.2-30.9 | <.001 |
| Anthacyclines | 11.5% | 5.3-19.8% | .964 |
| Topotecan | 7.2% | 2.8-13.3% | .002 |

\*significance for the “drug” variable in regression analysis
calculations were done with adjustment for response assessment and administration of platinum agents

### Progression-free survival and overall survival analysis

We attempted comparative analysis to assess an impact of various factors on PFS. Only the trials with reported median PFS were analyzed, so 104 trials with 5081 enrolled patients were included. Among them, 922 and 4159 patients were treated with platinum-based and non-platinum chemotherapy (list is available as a supplement for this article). Log-transformed medians PFS were analyzed. The results are presented in Table 5, administration of platinum agents was the only factor significantly impacting the median PFS. Under the model assumptions patients treated with platinum-based chemotherapy were predicted to have median PFS 2 month longer compared to non-platinum counterparts (p <0.001). Multivariate linear regression confirmed the independent impact of platinum agents on median PFS after adjustment for other factors (as neither evaluated factor was significant in univariable analysis it was decided to include all the factors without lost data), results are presented in Table 6.

**Table 5.** Linear regression univariable analysis of factors impacting the probability of response

| Variable | Intercept ($\beta_0$, 95% CI) | Estimate ($\beta_1$, 95% CI) | p | R<sup>2</sup> |
| --- | --- | --- | --- | --- |
| Platinum agents | 1.219 (1.157-1.281) | .462 (.316-.608) | **<.001** | 26.8% |
| CA-125 response | 1.302 (1.217-1.388) | .003 (-.133-.140) | .964 | .0% |
| Taxanes | 1.281 (1.209-1.352) | .154 (-.031-.339) | .103 | .0% |
| Etoposide | 1.295 (1.227-1.363) | .183 (-.136-.502) | .258 | .0% |
| Gemcitabine | 1.301 (1.227-1.374) | .01 (-.162-.187) | .889 | .0% |
| Anthracyclines | 1.320 (1.241-1.399) | -.058 (-.206-.089) | .436 | .0% |
| Topotecan | 1.311 (1.236-1.385) | -.035 (-.201-.130)) | .672 | .0% |
| Serous histology<sup>1</sup> | .821 (.125-1.517) | .007 (-.002-.016)) | .145 | .0% |
| Age<sup>1</sup> | .943 (-.373-2.260) | .006 (-.016-.028) | .543 | .0% |
| Prior therapy<sup>2</sup> | 1.456 (1.288-1.624) | -.070 (-.152-.0130) | .097 | .0% |

<sup>1</sup>per percent or unit; <sup>2</sup>per median of lines; data presented as mixed-effect model after log-transformation of median PFS

**Table 6.** Multiple linear model of variables impacting the PFS

| Variable | Estimate | 95% CI | Standard error | p |
| --- | --- | --- | --- | --- |
| Intercept ($\beta_0$) | 1.167 | 1.011-1.323 | .079 | **<.001** |
| Platinum agents | .507 | .342-.672 | .008 | **<.001** |
| CA-125 response | -.017 | .142-.108 | .063 | .789 |
| Taxanes | .138 | -.074-.351 | .107 | .199 |
| Etoposide | .040 | -.276-.356 | .159 | .804 |
| Anthracyclines | .075 | -.111-.261 | .093 | .424 |
| Topotecan | .099 | -.104-.302 | .102 | .334 |

data presented as mixed-effect model after log-transformation of median PFS
Adjusted $R^2 = .264$ , F-statistic: 6.282, $p < 0.0001$

**Table 7.** Linear regression univariable analysis of factors impacting the probability of response

| Variable | Intercept ($\beta_0$, 95% CI) | Estimate ($\beta_1$, 95% CI) | p | $R^2$ |
| --- | --- | --- | --- | --- |
| Platinum agents | 2.439 (2.381-2.498) | -.027 (-.168-.113) | .700 | .0% |
| Taxanes | 2.415 (2.356-2.473) | .115 (-.025-.255) | .108 | .0% |
| Etoposide | 2.439 (2.384-2.493) | -.098 (-.348-.152) | .473 | .0% |
| Gemcitabine | 2.436 (2.377-2.495) | -.011 (-.154-.132) | .880 | .0% |
| Anthracyclines | 2.431 (2.369-2.494) | .013 (-.109-.135) | .836 | .0% |
| Topotecan | 2.442 (2.383-2.502) | -.043 (-.182-.094) | .532 | .0% |
| Serous histology<sup>1</sup> | 2.029 (1.566-2.493) | .006 (-.001-.012) | .078 | .0% |
| Age<sup>1</sup> | 2.932 (1.902-3.962) | -.008 (-.026-.009) | .333 | .0% |
| **Prior therapy<sup>2</sup>** | **2.630 (2.501-2.760)** | **-.094 (-.154- -.033)** | **.003** | **10.5%** |

<sup>1</sup>per percent or unit; <sup>2</sup>per median of lines; data presented as mixed-effect model after log-transformation of median OS

Using the same methodology we analyzed OS of patients, 99 trials with 5089 patients were included in the analysis, 902 and 5089 patients were treated with platinum and non-platinum chemotherapy. The only factor significantly affecting OS was median of prior anticancer regimens. As no therapeutic option significantly impacted the results we did not conducted multivariate analysis of OS.

## Discussion

In recurrent platinum-resistant or platinum-refractory EOC sequential single-agent non-platinum chemotherapy is still considered to be a standard of care. “Binary classification” of recurrent disease is still endorsed by many local and international guidelines which is justified by toxicity of platinum agents reducing quality of life due to acute and long-term toxicity^1^. Considering the palliative goals of therapy for PROC patients, indeed it may be reasonable to choose less aggressive approaches for patients who would not response to further platinum.

However, the current paradigm of ineffective platinum chemotherapy for patients with early-relapsed ovarian cancer is based on the results of a few clinical trials conducted in early 1990-th: in 1989 Blackledge et al. published analysis of 5 phase II trials (n = 93) and reported inverse correlation of response to platinum-based chemotherapy and time since last line of platinum^15^. In patients with PFI <6 months RR was 10% compared to 61% in patients with PFI above with threshold. Larger trial published by Eisenhauer et al. (n = 273) shown the similar results^16^. Mentioned trials share common flaws: small sample sizes; absence of control groups and treatment with suboptimal regimens. Nevertheless, this led to a paradigm of non-platinum chemotherapy as the preferred treatment option for ovarian cancer patients with so-called PROC. This hypothesis was never addressed in large-scale clinical trials. The only randomized trial in CARTHAXY trial enrolled patients with ovarian cancer recurrence within ≤6 months after first- or second-line treatment including a platinum and taxane^17^. Patients were randomized in 1:1:1 ratio in weekly paclitaxel 80 mg/m2 arm as monotherapy or in combination with carboplatin AUC5 every 4 weeks or topotecan 3 mg/m2 weekly. The trial was clearly negative and shown no benefit of adding carboplatin or topotecan to paclitaxel monotherapy in PROC setting: rates of objective response and survival were similar across groups. RR were 35%, 37% И 39% in paclitaxel, paclitaxel/carboplatin and paclitaxel/topotecan groups respectively; median PFS – 3.7, 4.8 and 5.4 months (p = 0.46)^17^. However, there were several flaws in its design, as suboptimal dose of carboplatin, immediate retreatment with the very same regimen and unrealistic statistical assumptions. One should note, that RR in this trial was far lower than in other trials evaluated efficacy of paclitaxel/carboplatin retreatment in PROC setting^9,10^ and more than 50% of patients in this trial were platinum-refractory. The latter might influence the trial results and predict negative results of the trial.

The results of this meta-analysis illustrates that patients with PROC can derive significant benefit from reintroduction of platinum-based chemotherapy in terms of RR and PFS. Pooled response rate analysis was markedly higher in patients treated with platinum-based chemotherapy (37.6% ([95% CI 32.3-43.02]) and 16.37% [95% CI 14.11-18.77]) and these differences were statistically significant in multivariate meta-regression model adjusted for response assessment modality and use of various non-platinum agents (p < 0.001). Various sensitivity analyses confirmed the significance of platinum compounds as a response predictor.

Objective RR cannot be considered the ultimate treatment goal for ovarian cancer patients, however the current paradigm of platinum drugs inefficacy in PROC is based on very this endpoint. Furthermore, we attempted to assess an impact of platinum on progression-free survival. Analysis of log-transformed PFS medians confirmed significant advantages of platinum agents in patients with so-called “platinum resistant” ovarian cancer of more than 2 months (p <0.001) adjusted for other variables. We did not demonstrate any differences in OS related to platinum agents; however this endpoint is arguably far more dependent on previous and further anticancer therapy compared to RR and PFS.

For this meta-analysis we used acrsine transformation of raw proportions for better approximation to the normal distribution. Albeit, this method is not as popular as Freeman– Tukey (FT) double-arcsine transformation and logit-transformation it was chosen due to two main advantages. Compared to FT transformation arcsin transformed data may be easily back-transformed to raw proportion which is quite complicated for FT method and logit-transformation is impossible for zero event counts studies without adding some value to these trials which have an impact on the overall analysis results. After the main analysis we evaluated other transformation options as well and revealed the consistency of the results across all the transformation options.

We excluded all the trials involving targeted agents, mainly antiangiogenic drugs and PARP-inhibitors. Bevacizumab is often considered a standard of care for patients with PROC. However, we intentionally excluded trials with targeted agents as these may distort the results and introduce several biases. The design of our data analysis did not address the issue of dosing of various chemotherapeutic drugs, dose intensity and impact of these factors on outcomes as well. We were not able to investigate other potential time cut-offs as potential predictors of resistance to platinum agents. These are other limitations of this meta-analysis.

Our data analysis included more than 6000 patients and shown that patients with so-called “platinum-resistant ovarian cancer” benefit from platinum-based chemotherapy in terms of RR and PFS. This data looks quite encouraging and provide additional arguments in favor of platinum-based chemotherapy. On the other hand, one should interpret these findings with caution because indirect comparison of retrospective data is a subject to multiple biases. First of all, one can assume that patients who received non-platinum treatment had worse performance status and this could have detrimental impact on patients’ survival. Again, this was a retrospective analysis of various trials and it did not considered discrepancies among them in terms of disease stage, number of prior therapy lines, genetic alterations and many other factors.

In 2019 ESMO-ESGO consensus was published with revolutionary proposal to drop the binary 6 months cutoff^18^. According to the consensus, the definition of platinum resistance should be therapy-oriented and only patients who have progressed while receiving platinum-based chemotherapy or experienced a symptomatic relapse soon after the end of the last platinum-based chemotherapy to be considered platinum-resistant. Our results support recent ESMO-ESGO consensus on treatment of recurrent ovarian cancer and confirms platinum benefits in so-called “platinum-resistant” relapsed ovarian cancer.

## Conclusion

This systematic review provides an important signal that many patients with so-called “platinum-resistant ovarian cancer” are in fact at least sensitive to platinum agents. In conclusion, this systematic review shows that patients with ‘platinum-resistant’ ovarian carcinoma may derive significant benefit from reintroduction of platinum agents. This hypothesis should be confirmed in further randomized trials – one of such trials is currently ongoing (<u>NCT04055038</u>).

## Supporting information

Tab S1 - a trials data set

Figure S1 - Meta-proportional forest plot for all trials included in the analysis

## Data Availability

All data produced in the present study are available upon reasonable request to the authors

## Authors contribution

**Rumyantsev Alexey**: Writing and Editing, Conceptualization, Methodology, Investigation, Formal Analysis; **Tyulyandina Alexandra**: Investigation, Data Curation; **Fedyanin Mikhail**: Formal analysis; **Pokataev Ilya**: Formal analysis; **Glazkova Elena**: Investigation, Reviewing; **Tjulandin Sergey**: Supervision.

## Supplements

1. Figure S1 – Meta-proportional forest plot for all trials included in the analysis

2. Tab S1 – a trials data set

