## Supplementary material for "Platinum vs non-platinum chemotherapy for platinum-resistant ovarian cancer: a systematic review and meta-analysis": Tab S1 - a trials data set

| N | Author | Year | Ref |
| --- | --- | --- | --- |
| 1 | Elkas J | 2003 | ^1^ |
| 2 | Hoekstra A | 2009 | ^2^ |
| 3 | Denschlag D | 2004 | ^3^ |
| 4 | Rose P | 2003 | ^4^ |
| 5 | Kakolyris S | 2001 | ^5^ |
| 6 | Sharma R | 2011 | ^6^ |
| 7 | Polyzos A | 2000 | ^7^ |
| 8 | Havrilevsky L | 2003 | ^8^ |
| 9 | Tomita Y | 2014 | ^9^ |
| 10 | de Jongh F | 2002 | ^10^ |
| 11 | Vergote I | 2009 | ^11^ |
| 12 | Vergote I | 2009 | ^11^ |
| 13 | Levy T | 2004 | ^12^ |
| 14 | Shawky H | 2014 | ^13^ |
| 15 | Handolias D | 2016 | ^14^ |
| 16 | Downs L | 2008 | ^15^ |
| 17 | Ichikawa R | 2014 | ^16^ |
| 18 | Vandenput I | 2007 | ^17^ |
| 19 | Brown J | 2003 | ^18^ |
| 20 | Hu C | 2015 | ^19^ |
| 21 | Mitchell S | 2005 | ^20^ |
| 22 | Wilailak S | 2004 | ^21^ |
| 23 | Recchia F | 2007 | ^22^ |
| 24 | Kita T | 2004 | ^23^ |
| 25 | Rosa D | 2008 | ^24^ |
| 26 | Nagourney R | 2003 | ^25^ |
| 27 | Nicoletto M | 2006 | ^26^ |
| 28 | Sundar S | 2004 | ^27^ |
| 29 | Piura B | 2005 | ^28^ |
| 30 | Zanotti K | 2000 | ^29^ |
| 31 | Raspagliesi F | 2004 | ^30^ |
| 32 | Verborg W | 2008 | ^31^ |
| 33 | O'Malley D | 2005 | ^32^ |
| 34 | Marchetti C | 2017 | ^33^ |
| 35 | Meyer T | 2001 | ^34^ |
| 36 | Gorumlu G | 2008 | ^35^ |
| 37 | Malik I | 2001 | ^36^ |
| 38 | Harnett P | 2007 | ^37^ |
| 39 | Tsubamoto H | 2013 | ^38^ |
| 40 | Anand A | 2004 | ^39^ |
| 41 | Adam J | 2017 | ^40^ |
| 42 | Yuan S | 2013 | ^41^ |
| 43 | Bruchim I | 2016 | ^42^ |
| 44 | Stein S | 2013 | ^43^ |
| 45 | Shamsunder S | 2000 | ^44^ |
| 46 | Niwa Y | 2003 | ^45^ |
| 47 | Ferrandina G | 2014 | ^46^ |
| 48 | Sharma R | 2009 | ^47^ |
| 49 | Safra T | 2006 | ^48^ |
| 50 | Vasey P | 2003 | ^49^ |
| 51 | Germano D | 2007 | ^50^ |
| 52 | Le T | 2008 | ^51^ |
| 53 | Alici S | 2003 | ^52^ |
| 54 | Chanpanitkitchot S | 2014 | ^53^ |
| 55 | Tewari D | 2004 | ^54^ |
| 56 | Karabulut B | 2005 | ^55^ |
| 57 | Ghamande S | 2003 | ^56^ |
| 58 | Rose P | 2000 | ^57^ |
| 59 | Kurzeder C | 2016 | ^58^ |
| 60 | Kurzeder C | 2016 | ^58^ |
| 61 | Dear R | 2010 | ^59^ |
| 62 | Gounaris I | 2016 | ^60^ |
| 63 | Arimoto T | 2012 | ^61^ |
| 64 | Linch M | 2008 | ^62^ |
| 65 | Yoshino K | 2012 | ^63^ |
| 66 | Kang H | 2009 | ^64^ |
| 67 | Kalykaki A | 2008 | ^65^ |
| 68 | Safra T | 2007 | ^66^ |
| 69 | Matsumoto K | 2006 | ^67^ |
| 70 | Kurzeder C | 2016 | ^68^ |
| 71 | van der Burg M | 2002 | ^69^ |
| 72 | Chou H | 2006 | ^70^ |
| 73 | Markman M | 2000 | ^71^ |
| 74 | Poveda A | 2017 | ^72^ |
| 75 | Conteduca V | 2018 | ^73^ |
| 76 | Conteduca V | 2018 | ^73^ |
| 77 | Berkenblit A | 2004 | ^74^ |
| 78 | Markman M | 2003 | ^75^ |
| 79 | Bodurka D | 2003 | ^76^ |
| 80 | Pisano C | 2009 | ^77^ |
| 81 | Le T | 2006 | ^78^ |
| 82 | Rischin D | 2004 | ^79^ |
| 83 | Katsumata N | 2000 | ^80^ |
| 84 | Elshebeiny M | 2016 | ^81^ |
| 85 | Montazeri A | 2002 | ^82^ |
| 86 | Wong C | 2017 | ^83^ |
| 87 | McNeish I | 2014 | ^84^ |
| 88 | Wolf J | 2006 | ^85^ |
| 89 | Rodriguez M | 2001 | ^86^ |
| 90 | Pignata S | 2015 | ^87^ |
| 91 | Hensley M | 2012 | ^88^ |
| 92 | Song T | 2013 | ^89^ |
| 93 | Salah-Eldin M | 2012 | ^90^ |
| 94 | Pectasides D | 2004 | ^91^ |
| 95 | Morgan R | 2018 | ^92^ |
| 96 | Morgan R | 2018 | ^92^ |
| 97 | Ledermann J | 2010 | ^93^ |
| 98 | Ojeda Gonzales B | 2008 | ^94^ |
| 99 | D'Agostino G | 2003 | ^95^ |
| 100 | Vici P | 2013 | ^96^ |
| 101 | Gronlund B | 2002 | ^97^ |
| 102 | van der Burg M | 2013 | ^69^ |
| 103 | Cadron I | 2013 | ^98^ |
| 104 | Markman M | 2000 | ^99^ |
| 105 | Largillier R | 2007 | ^100^ |
| 106 | Largillier R | 2007 | ^100^ |
| 107 | Bozas G | 2007 | ^101^ |
| 108 | Coleman R | 2011 | ^102^ |
| 109 | Markman M | 2006 | ^103^ |
| 110 | De Geest K | 2010 | ^104^ |
| 111 | Strauss H | 2008 | ^105^ |
| 112 | Chekerov R | 2018 | ^106^ |
| 113 | Ray-Coquard I | 2009 | ^107^ |
| 114 | Miller D | 2009 | ^108^ |
| 115 | Prasad M | 2004 | ^109^ |
| 116 | Markman M | 2003 | ^110^ |
| 117 | Campos S | 2001 | ^111^ |
| 118 | Kucukoner M | 2012 | ^112^ |
| 119 | Lortholary A | 2012 | ^112^ |
| 120 | Bozkaya Y | 2017 | ^113^ |
| 121 | Naumann R | 2013 | ^114^ |
| 122 | Peled Y | 2013 | ^115^ |
| 123 | Pujade-Lauraine E | 2014 | ^116^ |
| 124 | Prasad M | 2004 | ^109^ |
| 125 | Meier W | 2009 | ^117^ |
| 126 | Coleman R | 2014 | ^118^ |
| 127 | Lortholary A | 2012 | ^119^ |
| 128 | Brewer C | 2006 | ^120^ |
| 129 | Rose P | 2003 | ^121^ |
| 130 | Vergote I | 2010 | ^122^ |
| 131 | Pujade-Lauraine E | 2014 | ^116^ |
| 132 | Calcagno M | 2009 | ^123^ |
| 133 | Pujade-Lauraine E | 2014 | ^116^ |
| 134 | Adams S | 2011 | ^124^ |
| 135 | Makhija S | 2010 | ^125^ |
| 136 | Clarke-Pearson D | 2001 | ^126^ |
| 137 | Takei Y | 2017 | ^127^ |
| 138 | Suprasert P | 2012 | ^128^ |
| 139 | Abushahin F | 2008 | ^129^ |
| 140 | Rose P | 2001 | ^130^ |
| 141 | Liu J | 2016 | ^131^ |
| 142 | Safra T | 2013 | ^132^ |
| 143 | Steppan I | 2009 | ^133^ |
| 144 | Ferrandina G | 2008 | ^134^ |
| 145 | Gordon A | 2000 | ^135^ |
| 146 | Hensley M | 2001 | ^136^ |
| 147 | Mutch D | 2007 | ^137^ |
| 148 | Mutch D | 2007 | ^137^ |
| 149 | Omura G | 2003 | ^138^ |
| 150 | Omura G | 2003 | ^138^ |
| 151 | Monk B | 2010 | ^139^ |
| 152 | Gordon A | 2001 | ^140^ |
| 153 | Gordon A | 2001 | ^140^ |
| 154 | Sehouli J | 2011 | ^141^ |
| 155 | Sehouli J | 2011 | ^141^ |
| 156 | Vergote I | 2009 | ^142^ |
| 157 | Colombo N | 2012 | ^143^ |

1.	Elkas JC, Holschneider CH, Katz B, et al. The use of continuous infusion topotecan in persistent and recurrent ovarian cancer. *International Journal of Gynecological Cancer*. 2003;13(2):138-141. doi:10.1046/j.1525-1438.2003.13020.x

2.	Hoekstra AV, Hurteau JA, Kirschner CV, Rodriguez GC. The combination of monthly carboplatin and weekly paclitaxel is highly active for the treatment of recurrent ovarian cancer. *Gynecologic Oncology*. 2009;115(3):377-381. doi:10.1016/j.ygyno.2009.08.021

3.	Denschlag D, Watermann D, Hörig K, Kissel C, Tempfer C, Gitsch G. Topotecan as a Continuous Infusion Over 14 Days in Recurrent Ovarian Cancer Patients. :3.

35.	Gorumlu G, Kucukzeybek Y, Kemal-Gul M, et al. Pegylated liposomal doxorubicin in heavily pretreated epithelial ovarian cancer patients. :4.

36.	Malik I. Altretamine is an effective palliative therapy of patients with recurrent epithelial ovarian cancer. *Jpn J Clin Oncol*. Published online 2001.

82.	Montazeri A, Culine S, Laguerre B, et al. Individual Adaptive Dosing of Topotecan in Ovarian Cancer. :7.
