## Supplementary figures and images for "Platinum vs non-platinum chemotherapy for platinum-resistant ovarian cancer: a systematic review and meta-analysis"

### Figure S1 - Meta-proportional forest plot for all trials included in the analysis

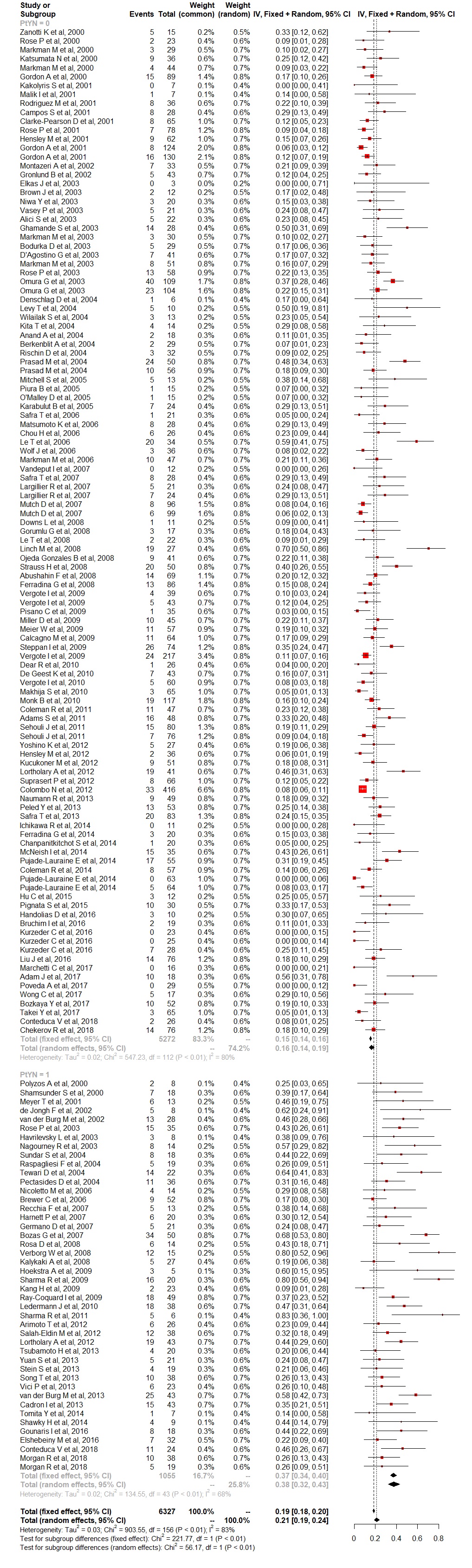
